# Three months of weekly rifapentine and isoniazid versus four months of rifampicin for tuberculosis infection: a randomised controlled trial

**DOI:** 10.1101/2025.08.18.25330309

**Authors:** Vicky L Chang, Qingbin Li, David Barnes, Anthony L. Byrne, Jin-Gun Cho, Sei Wei Foo, Hazel Goldberg, Timothy Gray, Zinta Harrington, Lydia M Rofail, Evan Ulbricht, Greg J Fox

**Affiliations:** Discipline of Medicine, Sydney Medical School, University of Sydney, including Concord, Nepean, Northern, Central (RPA), Westmead, and School of Rural Health Clinical Schools, NSW, Australia; St George Hospital, Gray St, Kogarah NSW 2217; Westmead Hospital, Corner Hawkesbury Road and Darcy Road, Westmead New South Wales 2145; University of Sydney, Faculty of Medicine and Health, NSW, Australia; Royal Prince Alfred Hospital, 50 Missenden Road, Camperdown NSW 2050; St Vincent Hospital, 390 Victoria Street, Darlinghurst New South Wales 2010; University of New South Wales, School of Clinical Medicine, Sydney NSW 2052; Concord Repatriation General Hospital, Hospital Road, Concord New South Wales 2139; Western Sydney University, Macarthur Clinical School, Sydney NSW 2560; Prince of Wales Hospital, L 3 High St E, Randwick New South Wales 2031; Canterbury Hospital, 575 Canterbury Road, Campsie New South Wales 2194; Liverpool Hospital, 1 Elizabeth Street, Liverpool New South Wales 2170; NSW TB program, NSW health; NHMRC Clinical Trials Centre, The Faculty of Medicine and Health, The University of Sydney, Level 6, Gloucester House, Royal Prince Alfred Hospital, 50 Missenden Road, Camperdown NSW 2050

## Abstract

**Background:** Treatment of tuberculosis infection (TBI) is a cornerstone of the WHO End TB Strategy. Two widely recommended short-course regimens—weekly isoniazid plus rifapentine for 12 weeks (3HP) and daily rifampicin for 16 weeks (4RIF)—have not been directly compared in a randomised trial. This study aimed to compare treatment completion between 3HP and 4RIF among individuals with TBI.

**Methods:** We conducted a multicentre, open-label, parallel-group, randomised controlled trial at seven tuberculosis clinics in Sydney, Australia, between July 1, 2019, and June 30, 2024. Participants of any age with TBI were randomised 1:1 using a computer-generated, site-stratified sequence to receive either 3HP (isoniazid 15 mg/kg [max 900 mg] plus rifapentine 900 mg weekly for 12 weeks) or 4RIF (rifampicin 10 mg/kg [max 600 mg], daily for 16 weeks). All doses were self-administered. Participants in the 3HP group received weekly SMS reminders; both groups received standard clinic follow-up. The primary outcome was treatment completion, defined as ingestion of ≥90% of prescribed doses, assessed at the end of treatment. Analyses followed the intention-to-treat principle, including all randomised participants. The trial is registered with the Australian New Zealand Clinical Trials Registry (ACTRN12618001672246) and is closed to new enrolments.

**Findings:** A total of 210 participants were enrolled (106 assigned to 3HP, 104 to 4RIF; 121 [57.6%] female, 89 [42.4%] male). Completion was higher in the 3HP group (90/106 [84.9%]) than the 4RIF group (68/104 [65.4%]; relative risk 1.30, 95% CI 1.10–1.53; p=0.002). Adverse events of any grade occurred in 26/106 (24.5%) in the 3HP group and 21/104 (20.2%) in the 4RIF group. No treatment-related deaths were reported.

**Interpretation:** The 3HP regimen, supported by SMS reminders, led to significantly higher treatment completion than 4RIF. These findings support broader implementation of 3HP in TBI programs to improve adherence and outcomes.

**Funding:** This study was funded by an Investigator Grant (#2007920) from the Australian National Health and Medical Research Council (NHMRC), with additional in-kind support from Sanofi Pharmaceuticals and the generous assistance of staff at the participating tuberculosis clinics.

## Background

Tuberculosis is a major global public health problem, affecting 10.6 million people in 2021 worldwide [1]. According to the World Health Organisation (WHO), approximately one-quarter of the global population has been infected with *M. tuberculosis* [1]. Healthy individuals can harbour TBI for many years, however, an estimated 5% of people infected with TB progress rapidly to active disease [2], leading to new infections among close contacts. Prevention of active TB disease by treatment of TBI, is therefore a critical component of the WHO End TB Strategy [3].

WHO has endorsed a number of alternative regimens for treatment of TBI, including six to nine months of daily isoniazid (6-9H) [4], 4 months of daily rifampicin (4RIF) [5, 6] 3–4 months daily rifampicin plus isoniazid [7] and 3-month weekly regimen of rifapentine plus isoniazid (3HP) [8]. An open-label randomised controlled trial demonstrated that 3HP was non-inferior to nine months of isoniazid (9H) [9], while having less hepatotoxicity and higher rates of treatment completion [9–12]. Similarly, the 4RIF regimen has also been shown to be non-inferior to isoniazid in both adults and children [13, 14]. With greater tolerability and shorter duration, rifamycin-based regimens have largely replaced isoniazid as standard therapy for treatment of TB infection that is presumed susceptible to first-line drugs. However, limited evidence is available to inform the decision about which rifamycin-based regimens should be offered to patients. Furthermore, the original phase 3 trial of 3HP regimen employed in-clinic observation of therapy, which is resource intensive and inconvenient for many. A subsequent Phase four clinical trial comparing self-administered and clinic-based therapy found higher rates of treatment completion in the clinic[15]. The one exception was the USA, in which treatment completion for self-administered therapy was lower compared with self-administration-with-reminders group, but within the 15% non-inferiority margin. Another study of two-way text messaging found no effect of weekly text messaging for daily self-administered treatment with rifampicin upon treatment adherence[16]. Therefore, further evidence is required to determine the optimal rifamycin-based regimen, and determine whether remote treatment adherence support can achieve high rates of adherence among people taking the 3HP regimen.

Therefore, we undertook a randomised controlled trial to compare treatment completion amongst patients with TBI assigned to the weekly 3HP using SMS monitoring, with patients assigned to daily self-administered 4RIF.

## Methods

### Study design and setting

A multi-centre, prospective, open label, phase four randomised controlled trial was conducted in seven chest clinics in Sydney, New South Wales (NSW), Australia. This network of publicly-funded tuberculosis clinics provides free treatment to residents of all ages regardless of immigration status. Chest clinics provide assessment and diagnosis for TBI for those at high risk of TB disease including contacts of people with TB disease and recent migrants.

### Study population and eligibility

The study population comprised patients aged two years and over attending participating clinics with a diagnosis of TBI. Participants were eligible if they were recommended to take treatment for TBI by a specialist Respiratory or Infectious Disease Physician. TBI was defined as a Tuberculin test (TST) of 10mm or greater, or positive on interferon gamma release assay (IGRA). People living with HIV were also eligible for treatment regardless of TB infection status, including those with a negative TST result, provided there was no evidence of active TB disease.. Eligible participants were required to have access to either a mobile or landline telephone. Exclusion criteria included patients who were currently pregnant confirmed by a urinary pregnancy test (BHCG), planning to become pregnant during the study period, breastfeeding, under consideration for solid organ transplantation, or with suspected TB infection resistance to either isoniazid or rifampicin. A structured questionnaire was completed at enrolment, including past medical history and demographic information.

Prior to randomisation, eligible patients underwent clinical assessment and chest radiography (CXR) to exclude TB disease, according to standard practice. Patients diagnosed with TB disease were treated according to routine clinical care and were not eligible for randomisation.

### Randomisation and blinding

Eligible participants were randomised in a one-to-one ratio using computer-generated sequences on REDCap. Owing to the inherent differences in treatment frequency and duration, participants and investigators were not blinded to group allocation.

### Intervention and comparator regimens

The intervention regimen comprised 3HP (intervention arm) given once weekly for 12 weeks. Dosing was based on weight (Supplementary Table S1) [17]. For participants who weighed more than 50kg, the once weekly rifapentine dose was 900mg. Isoniazid dosing (once weekly) was also based on age and weight. In the 3HP arm, participants weighing 60kg or more were given nine 100mg isoniazid tablets and three 300mg rifapentine tablets (a total of 12 tablets). In the 3HP group, adherence was supported through once-weekly SMS sent by clinic staff to participants’ mobile phones, or phone calls for those unable to use SMS. Participants were asked to respond to the message, otherwise a follow-up phone call was performed by a clinic nurse. The comparator regimen was daily self-administered rifampicin (4RIF) (weight-based dosing- 10mg/kg in adults (up to a maximum of 600 mg) and 15-20 mg/kg in infants & children-for a total 120-doses). Adherence to treatment in both groups was measured at each visit using pill counting by healthcare workers, pharmacy records of prescription fulfilment or via participant self-report.

### Safety monitoring

Prior to commencement of treatment for TBI, all participants had baseline liver function tests, and complete blood count performed. For women aged 15–45 years, a urinary pregnancy test (BHCG) was performed.

One month after commencement of treatment participants in both groups were assessed by the treating physician in accordance with routine clinical practice. This included a clinical assessment and repeat liver function tests to evaluate adherence and identify adverse events. Unscheduled clinic visits were available for participants who reported adverse events. At end of treatment participants were asked to complete an end-of-treatment questionnaire related to adverse events, their satisfaction about the treatment they received and the acceptability of the regimen. Participants in the 3HP group were also asked about their experience of SMS monitoring.

In the event of an adverse event the decision about whether to continue treatment was made by the treating practitioner in accordance with usual practice. If participants developed symptoms of an acute hypersensitivity reaction during treatment, considered an adverse event of special interest, they were asked to stop treatment. During the period of public health restrictions associated with the COVID-19 pandemic telehealth appointments were performed instead of face-face appointments for all visits.

### Study outcomes

The primary outcome was the proportion of participants completing treatment. This was defined as taking at least 90% of the required doses within no more than one month longer than the prescribed treatment period. For those in the 3HP arm this comprised at least 11 doses taken within 16 weeks of randomisation. For the 4RIF arm this comprised at least 108 of 120 doses taken within 5 months of randomisation.

Secondary outcomes included participant satisfaction, evaluated on the perceived convenience of the assigned regimen and the willingness to recommend it to others, and the proportion of participants who experienced at least one adverse event (AE) of any grade from the date of randomisation to the end of treatment. Adverse events were classified according to the National Cancer Institutes (US) Common Terminology Criteria for Adverse Events [18].

AEs were graded and managed by the treating clinician. Adverse events of special interest (AESI), and other clinical concerns were reviewed by site investigators.

### Monitoring for systemic drug reactions / hypersensitivity

Systemic drug reactions were considered to be an adverse event of special interest, with a reported incidence of 3.5% of among participants taking rifapentine [19]. If a participant developed this adverse event, then further tests were performed to further characterise the immunological nature of the adverse event.

A systemic drug reaction was defined according to established criteria [19]: (1) one of hypotension (systolic blood pressure <90 mm Hg), urticaria (hives), angioedema, acute bronchospasm, or conjunctivitis (red eyes); and (2) at least four of the following symptoms occurring concurrently (at least one of which had to be grade two or higher): weakness, fatigue, nausea, vomiting, headache, fever, aches, sweats, dizziness, shortness of breath, flushing, or chills.

### Sample size and statistical analysis

Assuming a 74% completion rate for the 4-month rifampicin regimen [12] and an 85% completion rate for the 3-month regimen delivered by in-person direct observation in the initial phase 3 trial of 3HP [21], we expected treatment completion would be 87% in the 3HP arm [22]. Allowing for up to 10% loss to follow-up, we estimated that 210 participants (105 per arm) would be required to achieve 80% power to detect superiority at a one-sided significance level of 0.05, without accounting for clustering.

In accordance with the pre-specified statistical analysis plan, the primary outcome of treatment completion rate was evaluated in the intention to treat (ITT) population, including all randomised participants regardless of treatment initiation, completion, or adherence. A binary regression model with robust standard errors was used to estimate the relative risk (RR). Secondary outcomes included adverse events and patient satisfaction, both assessed in the ITT population. The proportion of patients experiencing one or more adverse events was analyzed using a binary regression model with robust variance estimators. Patient satisfaction was measured using Likert scale responses and analyzed using ordinal logistic regression models with robust standard errors to estimate odds ratio (OR). Treatment group allocation was the only covariate adjusted for in both primary and secondary analyses.

To assess the robustness of our findings we performed several adjusted and sensitivity analyses. First, for both the primary and patient satisfaction outcomes, additional covariates of age and sex were included to adjust for potential confounding. No other clinically important baseline imbalances were identified. Second, for the primary and adverse event outcomes, we conducted analyses among participants who received at least one dose of the study treatment. The same models used in the main analyses were applied to these adjusted and sensitivity analyses.

Additional details of the analytic methods are provided in the Supplementary Appendix. Although the sample size was estimated using a one-sided hypothesis to optimise efficiency based on the expected direction of effect, all outcomes were analysed using two-sided tests and reported with two-sided 95% CIs and p values to preserve analytical rigour, ensure transparency, and minimise the risk of selective reporting. All statistical analyses were conducted using Stata 18 software (StataCorp, 2023).

### Site monitoring and quality assurance

Site monitoring was conducted at each site within 6 weeks of commencing recruitment and every six months subsequently or as needed. Quality assurance of the investigational product was undertaken by clinical trials pharmacies at each site.

### Ethical issues

The trial was registered prospectively with the Australian and New Zealand Clinical Trials Registry (www.anzctr.org.au; ACTRN12618001672246). Ethical approval for the study was granted by the Sydney Local Health District Human Research Ethics Committee (HREC/17/RPAH/229). A Clinical Trials Notification from the Therapeutic Goods Administration (TGA) was obtained (CT-2019-CTN-01472-1) to enable the use of rifapentine which was not licensed by the TGA. The study sponsor was Sydney Local Health District.

Prior to providing consent, potential participants aged 18 years and over were given a Participant Information Statement. Written informed consent was obtained from participants aged 18 and over. For participants under 18 years of age, a parent or guardian was asked to sign the Participant Consent Form. Participants between 10 and 17 years of age were also asked to sign an assent form.

### Financial interests

The rifapentine used for this study was provided by Sanofi Aventis Australia Pty Ltd (Sanofi). Sanofi did not contribute financially to the conduct of the study and was not involved in the study design, data analysis or interpretation, or dissemination of findings.

## Results

Between July 2019 and July 2024, 210 participants were enrolled and randomised within seven chest clinics. Of these, 106 (50.5%) were allocated to the 3HP intervention arm, and 104 (49.5%) were allocated to the 4RIF comparator arm. Out of the 210 participants who were randomised (Figure 1), 21 (10%) were lost to follow-up after randomization prior to completing treatment. Among these the proportion lost to follow up was 9/106 (8.5%) in the 3HP arm and 12/104 (11.5%) in the 4RIF arm. Additionally, one participant in the 3HP arm did not commence on treatment post-randomization due to low platelet levels from baseline blood tests and one participant from the 4RIF arm did not commence on treatment because of pregnancy. Nine participants received medications but were lost to follow-up afterward, two in the 3HP arm and seven in the 4RIF arm. All randomised participants were included in the analysis regardless of treatment initiation, completion, or adherence. The demographic and clinical characteristics of the participants in the two groups are shown in Table 1. Most participants (189/2010; 90%) were overseas-born, with 168 (80%) born in Asia. Just under half of participants were male (97/210; 46.2%). The median age of participants was 35.5 years (interquartile range [IQR]: 28.2-45.8 years). One of the patients was known to be HIV positive, although HIV testing was not performed as a part of the study. Most of the participants 188/210 (89.5%) reported no medical co-morbidities.

**Figure 1.**
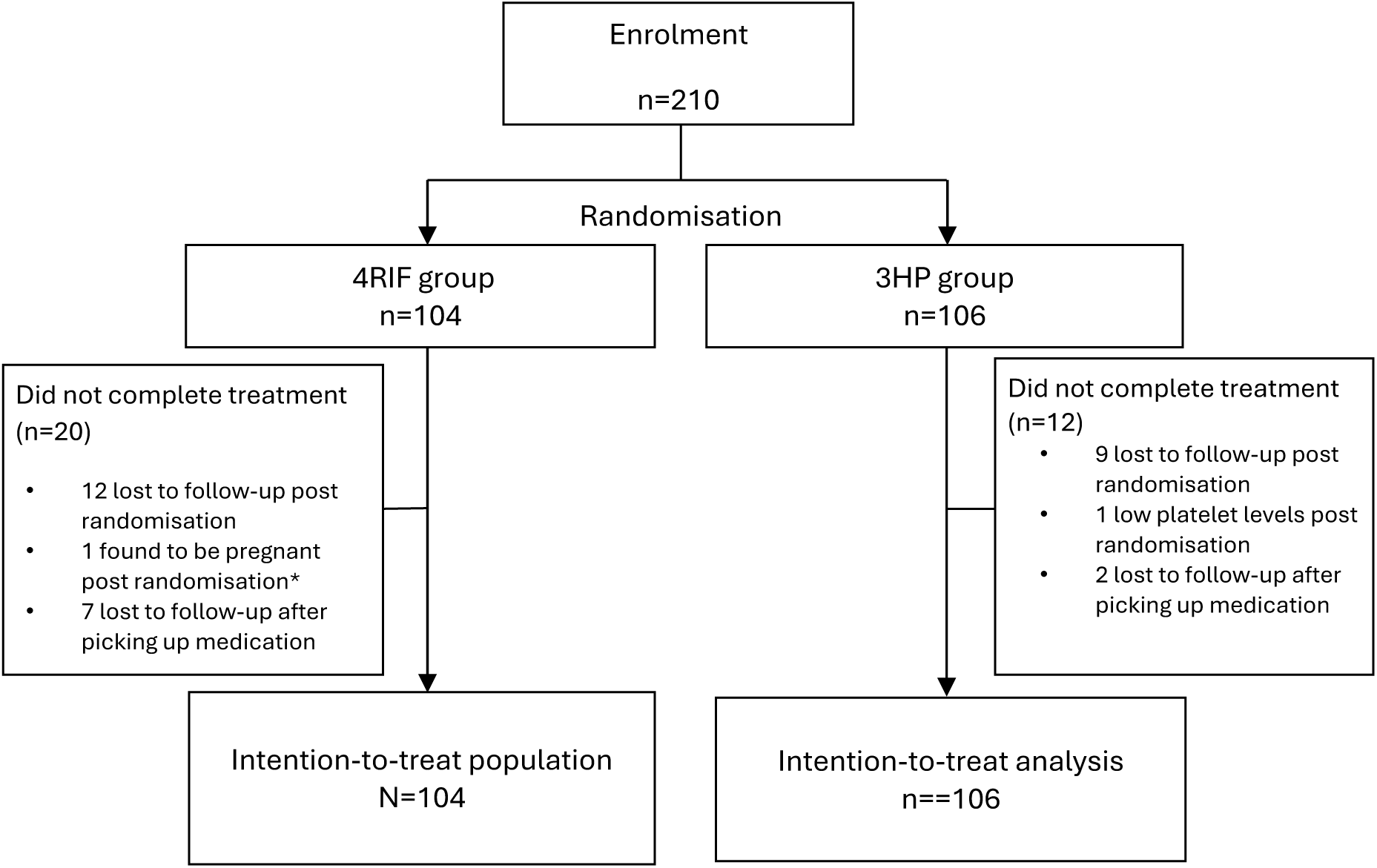
Consort diagram showing flow of participants through the study * One individual stopped treatment post-randomisation since the only post-randomisation exclusion based on a pre-randomisation criterion was pregnancy identified after randomisation. As pre-specified in the statistical analysis plan, the number of such cases was very small, making it unlikely that this exclusion would affect the balance achieved by randomisation or the overall study results. Therefore, in this study, the modified intention-to-treat (mITT) population was seen as identical to the ITT population, and no separate mITT analyses were conducted. Instead, sensitivity analyses were performed to assess the impact of excluding this case, as well as participants who never initiated study treatment.

**Table 1:**
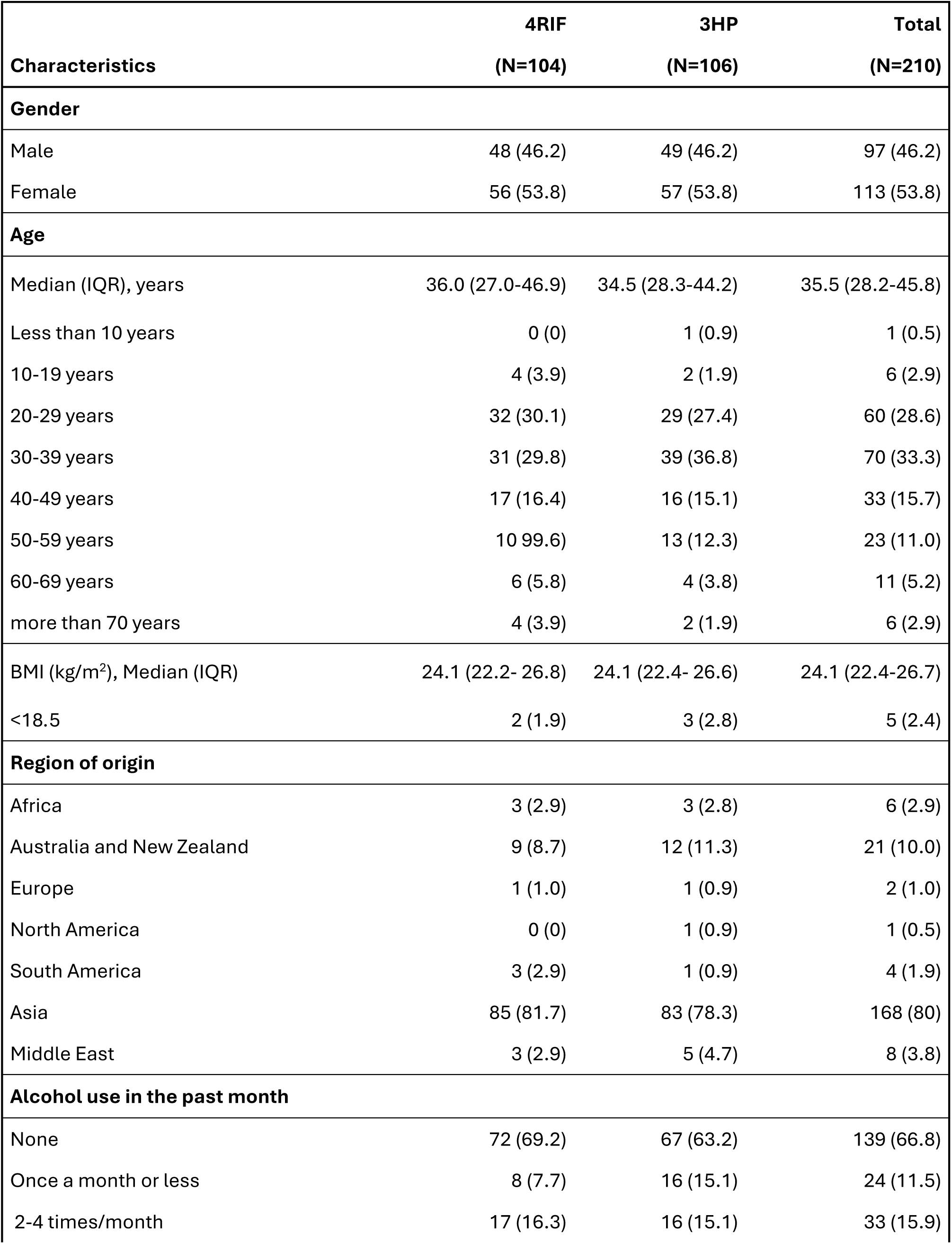
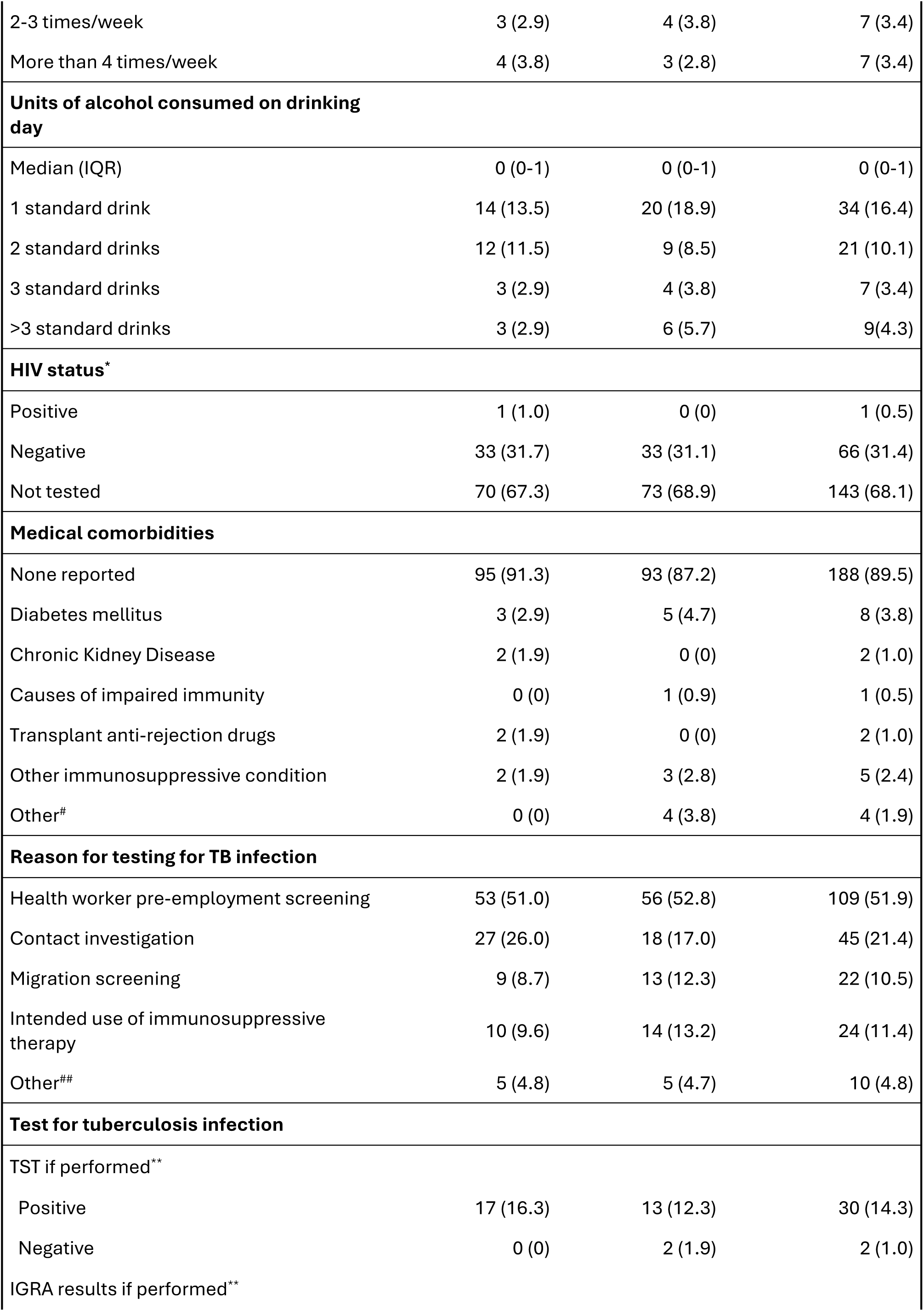
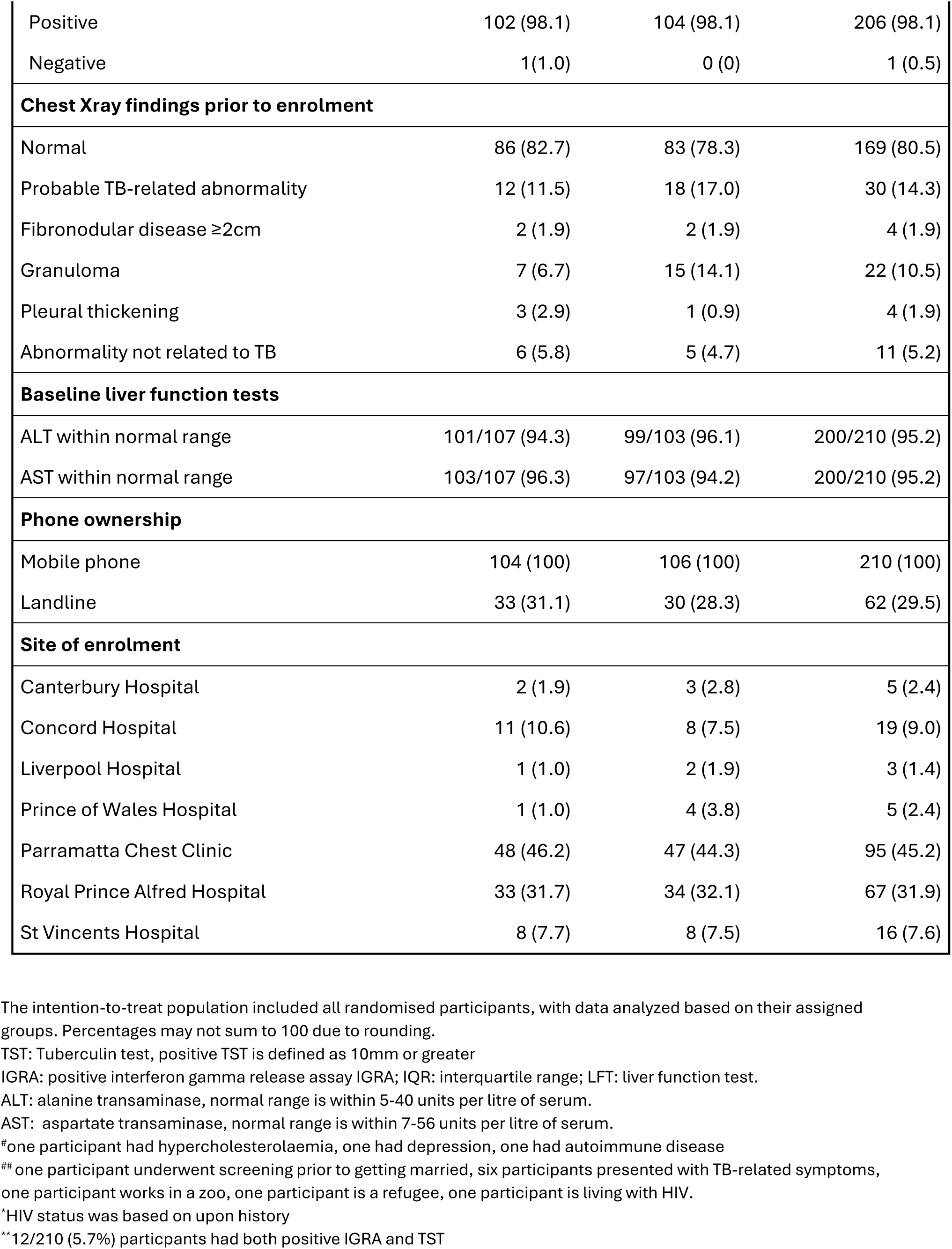
Characteristics of study participants, by group.

For the primary outcome 90 of 106 participants in the 3HP arm (84.9%) completed at least 90% of the required doses with no more than a one-month delay longer than the prescribed treatment period while 68 of 104 (65.4%) in the 4RIF arm completed treatment in this period. The relative risk of treatment completion for 3HP versus 4RIF was 1.30 (95% CI: 1.10–1.53, p = 0.002) indicating a 30% higher likelihood of completion among participants assigned to 3HP (Table 2). In a secondary adjusted analysis including age and gender, the results remained consistent with the primary unadjusted analysis (Supplementary Table S7).

**Table 2:** Primary and secondary outcomes in the intention-to-treat population, by group.

|  | 4RIF (n=104) | 3HP (n=106) | Effect Measure | Estimate (95%CI) | P value |
| --- | --- | --- | --- | --- | --- |
| **Primary outcome** |  |  |  |  |  |
| Proportion of participants completing treatment* | 68 (65.4%) | 90 (84.9%) | RR | 1.30 (1.10, 1.53) | 0.002 |
| **Secondary outcomes** |  |  |  |  |  |
| Perceived convenience of tuberculosis preventive therapy | -- | -- | OR | 2.66 (1.42, 4.98) | 0.002 |
| Likelihood of recommending the assigned regimen to others | -- | -- | OR | 3.86 (1.84, 8.07) | <0.0001 |
| Proportion of participants experiencing at least one adverse event from randomisation to the end of treatment | 21 (20.2%) | 26 (24.5%) | RR | 1.21 (0.73, 2.02) | 0.454 |

RR, relative risk. OR, odds ratio. CI, confidence interval. --" indicates responses were categorical (number and percentage), with details of the response presented in the appendix (Table S3). The database for the analysis was locked on January 31, 2025. All outcomes were evaluated in the intention-to-treat population during the overall study period. \*Defined as participants taking $\geq 90\%$ of prescribed doses within up to one month beyond the scheduled duration. 3HP participants had to complete $\geq 11$ of 12 doses within 16 weeks, while 4RIF participants needed $\geq 108$ of 120 doses within five months.

For the secondary outcomes in the intention-to-treat population, 159/210 (75.7%) of participants felt their tuberculosis preventive treatment (TPT) regimen was convenient or very convenient. A higher proportion of participants in the 3HP arm (43.4%) reported the regimen as very convenient compared to those in the 4RIF arm (19.2%). In the 3HP arm 81.1% participants stated they would recommend their assigned treatment to others and 67 of 104 (64.4%) in 4RIF arm (Supplementary Table S3, Figures S3, S4). Using ordinal logistic regression participants receiving the 3HP regimen were more likely to perceive the treatment as convenient (OR = 2.66, 95% CI: 1.42–4.98, p = 0.002) and to recommend it to others (OR = 3.86, 95% CI: 1.84–8.07, p < 0.0001) compared to those receiving the 4RIF regimen (Table 2). These findings remained consistent after adjusting for age and gender (see Supplementary Table S8).

In the 3HP group 82 of 106 (77.4%) responded to the SMS on all 12 weeks of the treatment. Among this group 83 of 106 (78.3%) found the SMS monitoring very helpful and 74 of 106 (69.8%) participants indicated they would choose SMS monitoring for TPT in the future (Supplementary Table S4, Figure S5).

### Adverse events

In the intention-to-treat population 26 of 106 (24.5%) in the 3HP arm and 21 of 104 (20.2%) in the 4RIF arm reported one or more adverse event. The relative risk of reporting at least one adverse event was 1.21 (95% CI: 0.73–2.02; p = 0.454), indicating no significant difference in the incidence of adverse events between the two treatment arms (Table 2).

Most adverse events were grade 1 or 2 (Table 3, Supplementary Table S5). More than one adverse event was reported by 4 of the 26 (15.4%) participants in the 3HP arm. There was one grade 3 adverse event that was classified as a systemic hypersensitivity reaction, an adverse event of special interest. The participant was taking rifampicin and presented to an emergency department with angioedema, fatigue, nausea, vomiting and dizziness a few hours after taking the dose for the first time. The participant was given intramuscular adrenaline, treated with hydrocortisone, went home after a period of observation and made a full recovery.

**Table 3:** Adverse events occurring from randomisation to the end of treatment in the intention-to-treat population, by group.

| Event | 4RIF<br>(n=104) | 3HP<br>(n=106) | Relative Risk<br>(95% CI) |
| --- | --- | --- | --- |
| **Participants with one or more adverse events** |  |  |  |
| Any (one or more) | 21 (20.2%) | 26 (24.5%) | 1.21 (0.73, 2.02) |
| Grades 3 and 4 adverse events* | 1 (1.0%) | 0 (0) | -- |
| Death | 0 (0) | 0 (0) | -- |
| **Permanent discontinuation of study drug because of adverse event** |  |  |  |
| Total | 12 (11.5%) | 4 (3.8%) | -- |
| Grades 3 and 4 adverse events* | 1 (1.0%) | 0 (0) | -- |
| **Adverse events of special interest** |  |  |  |
| Systemic drug reaction/hypersensitivity reaction<sup>#</sup> | 1 (1.0%) | 0 (0) | -- |

\*Defined as Grade 3: severe or medically significant but not immediately life-threatening; prolongation of hospitalisation indicated; disabling; limiting self-care activities of daily living; Grade 4: life-threatening consequences; urgent intervention indicated.
<sup>#</sup>The definition of a systemic drug reaction followed the definition used in the PREVENT TB trial [11]. The PREVENT Tuberculosis sub-study protocol team established two broad criteria for such a drug reaction; (1) hypotension (systolic blood pressure <90 mm Hg), urticaria (hives), angioedema, acute bronchospasm, or conjunctivitis (red eyes); and (2) at least 4 of the following symptoms occurring concurrently (>1 of which had to be grade 2 or higher): weakness, fatigue, nausea, vomiting, headache, fever, aches, sweats, dizziness, shortness of breath, flushing, or chills. AE that met either of the above criteria were termed systemic drug reactions (SDR).

Among the 47 participants who reported at least one adverse event (AE) 22 out of 26 patients (84.6%) in the 3HP arm and nine out of 21 patients (42.9%) in the 4RIF arm experienced no interruptions to their treatments with these events. However four out of 26 participants (15.4%) in the 3HP arm and 9 out of 21 patients (42.9%) in the 4RIF arm refused further treatment after experiencing an adverse event. Treating physicians discontinued treatment for three participants (2.9%) in the 4RIF arm, and none of these participants were offered an alternative regimen (Table 3, Supplementary Table S6).

Sensitivity analyses showed that the estimates for the primary outcome and AE outcome were consistent between the main analyses and among participants who received at least one dose of the study regimen (Supplementary Table S9).

## Discussion

This pragmatic, open-label, randomised controlled trial compared treatment completion and tolerability of three months of weekly rifapentine-isoniazid with SMS monitoring to four months of daily self-administered rifampicin for the treatment of TB infection in Australia. The study found that significantly more participants completed 3HP (84.9%) than 4RIF (65.4%) within the required period. Additionally, a greater proportion of participants in the 3HP arm (43.4%) reported the regimen as very convenient, compared to 19.2% in the 4RIF arm.

Adherence rates in this comparison of 3HP and 4RIF were similar to those observed in the original Phase 3 trials of these regimens, which found a 74% completion rate with 4RIF [12] and an 85% completion rate with 3HP supported by in-person direct observation in the clinic [21]. Having a direct comparison between these two regimens lends greater confidence that treatment completion is more likely with the 3HP regimen. The greater adherence with 3HP is likely related to the weekly adherence reminders provided by healthcare workers. As the regimen is taken only once weekly, sending two-way SMS is more feasible than it would be for a daily regimen. Asynchronous messaging by nursing staff is a simple and low-cost approach which strengthens the relationship with the patient. In most clinics, this was done on the same day each week, minimising the time required and assisting with workflow. This approach has also been shown to improve antiretroviral treatment adherence for HIV patients [11]. Further supporting this approach, most participants in our study found 3HP to be highly acceptable. Together, this indicates that remote SMS monitoring is a practical alternative to direct observation for the weekly regimen. It is important to acknowledge that only participants in the 3HP group received adherence reminders. Only participants in the 3HP group received adherence reminders, which may have contributed to higher completion rates; however, daily reminders for the 4RIF group would not have been practical or scalable in routine programmatic settings.

Both regimens were generally well-tolerated. While one in four participants in both arms reported at least one adverse event during treatment, almost all were grade 1-2, and most participants were able to complete treatment successfully after reporting an adverse event. Only one participant experienced systemic drug reaction/hypersensitivity reaction, in the rifampicin group. This is consistent with recent individual patient data meta-analyses demonstrating that the treatment-related grade 3 or 4 adverse events for 3HP and 4RIF that led to permanent drug discontinuation were rare (0.8% and 2.2% respectively) [20].

The study had several limitations. We excluded patients who were pregnant and being worked up for solid organ transplantation. Only one of the participants included in the study was known to be HIV positive. This may limit the generalisability in these populations. Secondly, this study included a large number of healthcare workers undergoing pre-employment screening and patients intended for immunosuppressive therapy. This group may be more likely to adhere to treatment than those who have a lower health literacy. Nevertheless, our inclusion criteria for the study were broad, and those who were enrolled were reflective of those commonly receiving TPT in Australia.

Our study has showed that both 3HP and 4RIF are feasible, tolerable and acceptable options for the treatment of TBI. However, the significantly higher likelihood of treatment completion with 3HP (RR 1.30, 95% CI: 1.10–1.53, p = 0.002), coupled with a greater proportion of participants reporting the regimen as very convenient, indicates that 3HP is acceptable to most patients.

This study provides important evidence for policy-makers in Australia and comparable high-income settings where TPT is widely used. Increasing access to rifapentine will not only provide patients with more choice, but can contribute to improved rates of treatment completion. Newer combination tablets that include 300mg of rifapentine and 300mg of isoniazid will reduce the pill burden further, to just 3 tablets weekly from the 12 tablets that were used each week in this study [21]. Furthermore, increasing access to 3HP may also increase the proportion of patients willing to consider preventive therapy, improving the progression through the cascade of care for TB infection [22].

Further research is required to determine the relative costs of the 3HP and 4RIF regimens. A previous pragmatic trial from Victoria, Australia showed that self-administered 3HP therapy was less costly than that of self-administered 9H therapy [14]. The availability of rifapentine through the Global Drug Facility will ensure increased access to affordable therapy that will support its further scale-up [23]. Secondly, even shorter treatments are being investigated to further improve adherence. A recent trial found that one month of daily rifapentine plus isoniazid was non-inferior to daily isoniazid for 9 months for the prevention of tuberculosis in HIV-infected adults [24]. Ongoing research in looking for novel shorter and more patient friendly TPT options in different population group will be crucial to increase the uptake of TBI treatment.

In conclusion, this clinical trial found that 3HP with SMS support was associated with a significantly higher completion rate (84.9%) compared to 4RIF (65.7%) in a high-income setting. Expanded availability of short-course therapies for TBI treatment will play a crucial role in government efforts to promote preventive therapy for TB in Australia and other comparable settings.

## Data Availability

All data produced in the present study are available upon reasonable request to the authors.

## Supplementary Appendix

### Trial Investigators

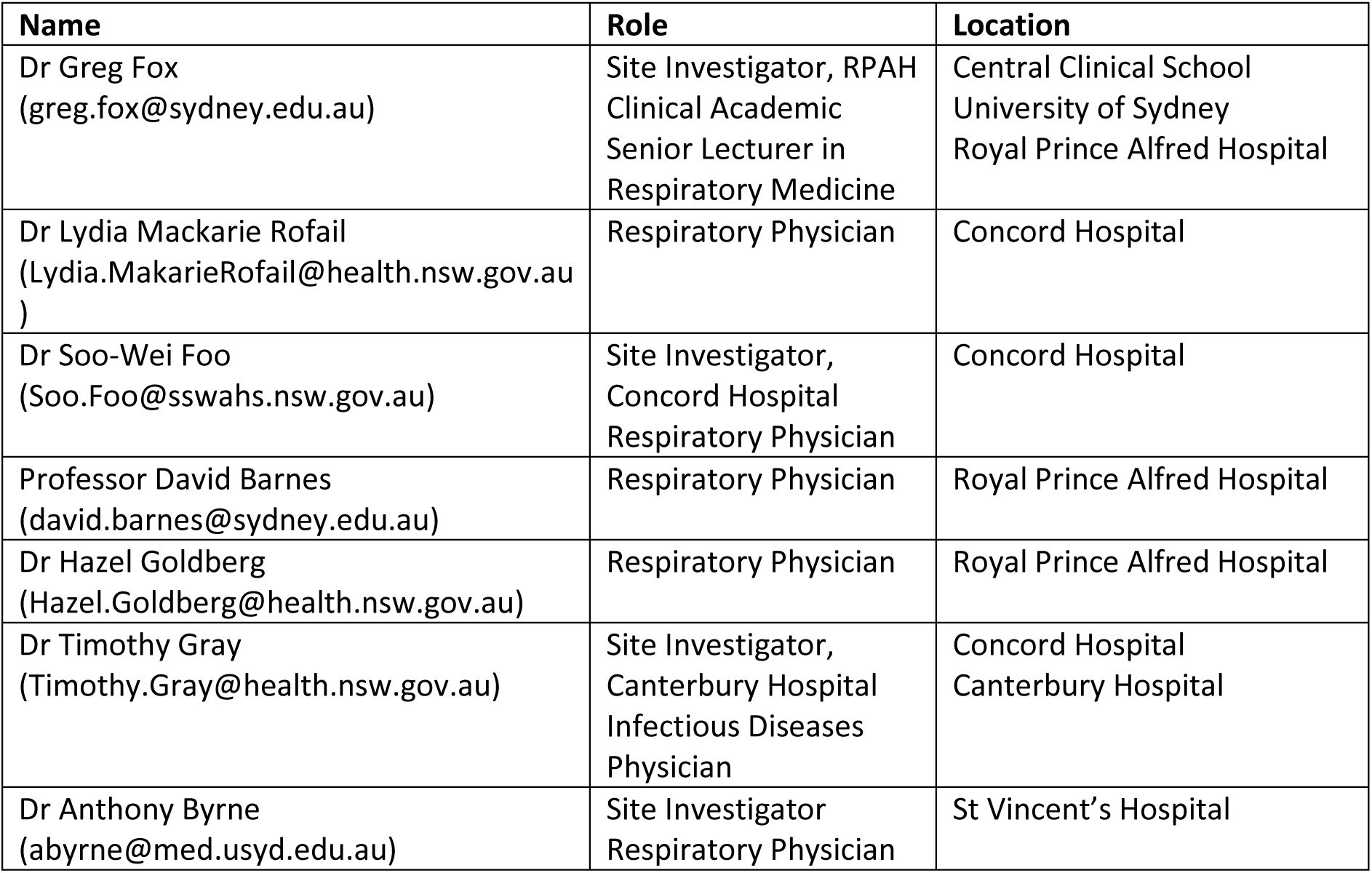

### Protocol Version Numbers

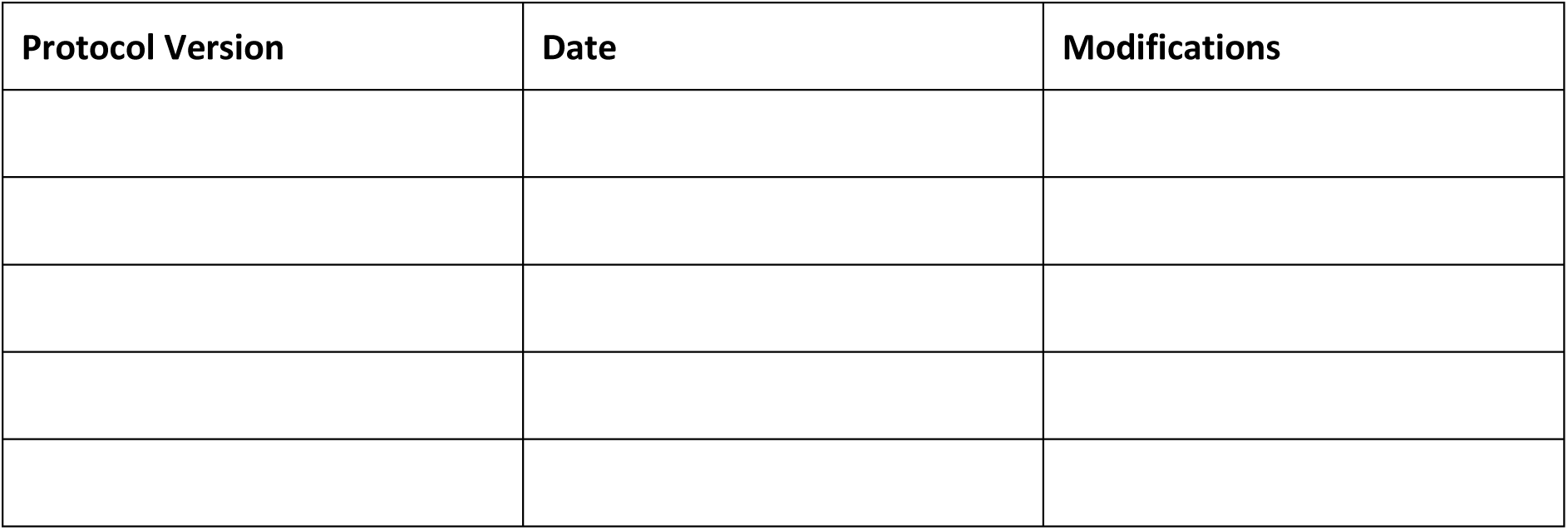

## Supplementary Methods

### Missing data

The primary outcome was analysed using the intention-to-treat (ITT) population. Participants who did not initiate study treatment were considered as not having completed the treatment. No other missing data were observed for the primary outcome; therefore, no imputation was necessary. Adverse events were also assessed within the ITT population. As adverse events were closely monitored throughout the study, there were no missing data for this outcome. Patient satisfaction was also evaluated in the ITT population. A small number of participants did not complete the end-of-study survey; hence a complete case analysis was used for this outcome. No missing data were observed in the adjusted or sensitivity analyses. Thus, no imputation methods were applied in this study.

### Multiplicity

No formal multiplicity adjustment was applied in this study. P-values were reported for the primary outcome (treatment completion rates), all pre-specified secondary outcomes, and key safety outcomes, as these were the main focus of the analysis. Where multiple comparisons were performed, the lack of formal adjustment for multiplicity was explicitly noted, and results were interpreted with caution.

### Analysis populations

Only the ITT population was primarily used in this study, which included all randomised participants regardless of treatment initiation, completion, or adherence. The ITT population was used for evaluating treatment completion rates, AEs, and patient satisfaction outcomes. No analyses were conducted using the modified Intention to Treat (mITT) population, as the only post randomisation exclusion based on a pre-randomisation criterion was pregnancy identified after randomisation. Since there was only one such case in the study, the ITT and mITT populations were effectively identical. Therefore, only the ITT population was used, with a footnote provided to document this situation.

### Statistical modelling

Both the primary outcome and the adverse event outcome were analyzed using binary regression models with a robust variance estimator. Model fit was assessed using the Akaike Information Criterion (AIC) and Bayesian Information Criterion (BIC), with lower values indicating a better fit. Additionally, plots of Pearson residuals versus predicted values were evaluated to identify potential model misfit.

For patient satisfaction outcomes, ordinal logistic regression models with robust standard errors were used. The proportional odds assumption was assessed using the Brant test.

To confirm the robustness of the results for both the primary outcome and patient satisfaction outcomes, additional covariates including age and sex were included in the models to adjust for potential confounding. Based on an assessment of baseline characteristics, no significant imbalance was observed, suggesting minimal confounding; therefore, no further covariates were adjusted for.

Two sensitivity analyses were conducted to assess the robustness of the study findings. For the primary outcome and AE outcome, the same main analyses were performed among participants who received at least one dose of the assigned study treatment. This was done because a small number of participants in each group did not initiate treatment, which could have influenced the results. By restricting the analysis to those who started treatment, we aimed to evaluate whether these two findings remained consistent under this scenario.

A crude intraclass correlation coefficient (ICC) was calculated based on the study data to confirm whether adjusting for clustering effects in the models was necessary.

## Supplementary Results

### Model fit and assumption check

Model fit was reasonable for all models, so no alternative models were used. The Brant test results for the two satisfaction outcomes both showed p-values greater than 0.05, indicating that the proportional odds assumptions were not violated. As a result, there was no need to use a generalized ordered logistic model.

### Adjusted models

For the primary outcome, the adjusted relative risk, accounting for age and sex, closely aligned with the unadjusted results, demonstrating the robustness of the findings (Table S7). For the patient satisfaction outcome, the adjusted odds ratios, also accounting for age and sex, remained consistent with the unadjusted results (Table S8).

### Sensitivity analyses

The estimates for the primary outcome and AE outcome were consistent between the main analyses and among participants who received at least one dose of the study treatment (Table S9). This suggests that the exclusion of participants who did not initiate treatment had minimal impact on the overall findings. The robustness of the results across these populations supports the reliability of the study conclusions.

### Intraclass correlation coefficient

The crude intraclass correlation coefficient calculated from the study data was 2.14 × 10⁻³⁴, effectively zero. This confirmed that there was no need to adjust for clustering in the analysis. It also validated that the omission of clustering adjustment in the sample size estimation was appropriate and did not affect the statistical power of the study.

## Supplementary Figures

**Figure S1.**
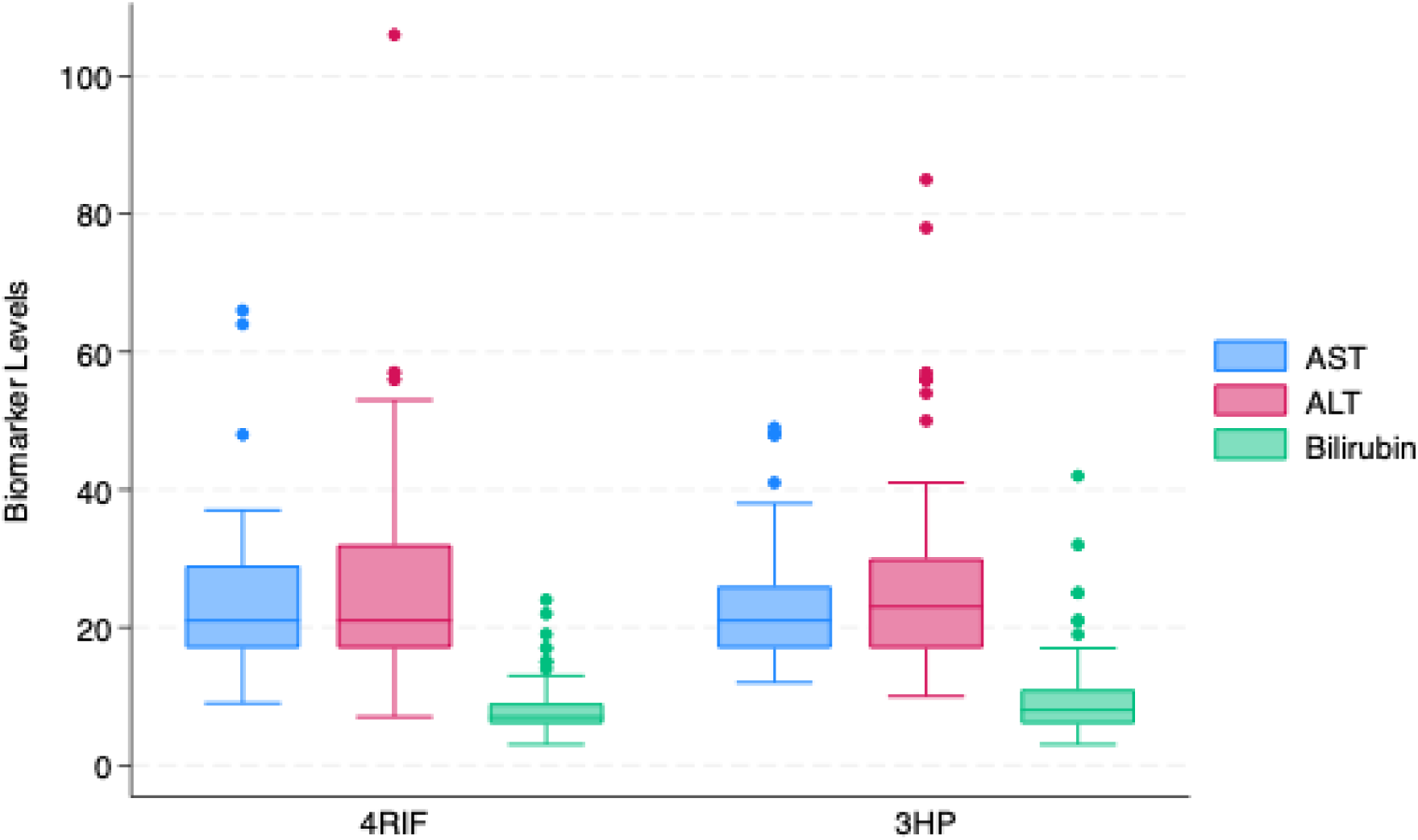
Boxplot of baseline liver function by group AST: Aspartate transaminase, units per litre; ALT: Alanine transaminase, units per litre; Bilirubin: micromoles per litre.

**Figure S2.**
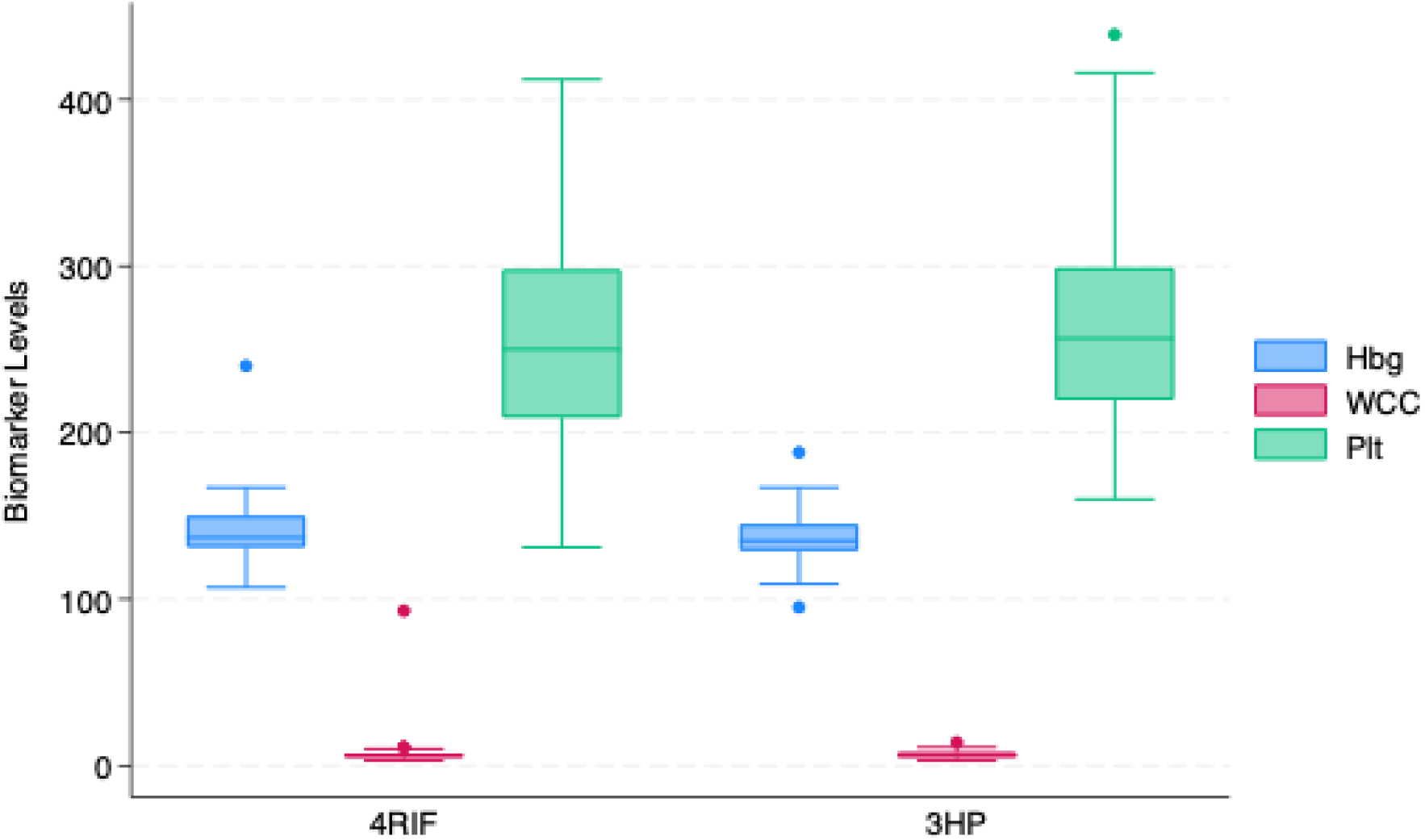
Boxplot of baseline haematological parameters by group Hbg: Haemoglobin, grams per litre; WCC: White Cell Count, cells ×10⁹/L; Plt: Platelet Count, cells ×10⁹/L.

**Figure S3.**
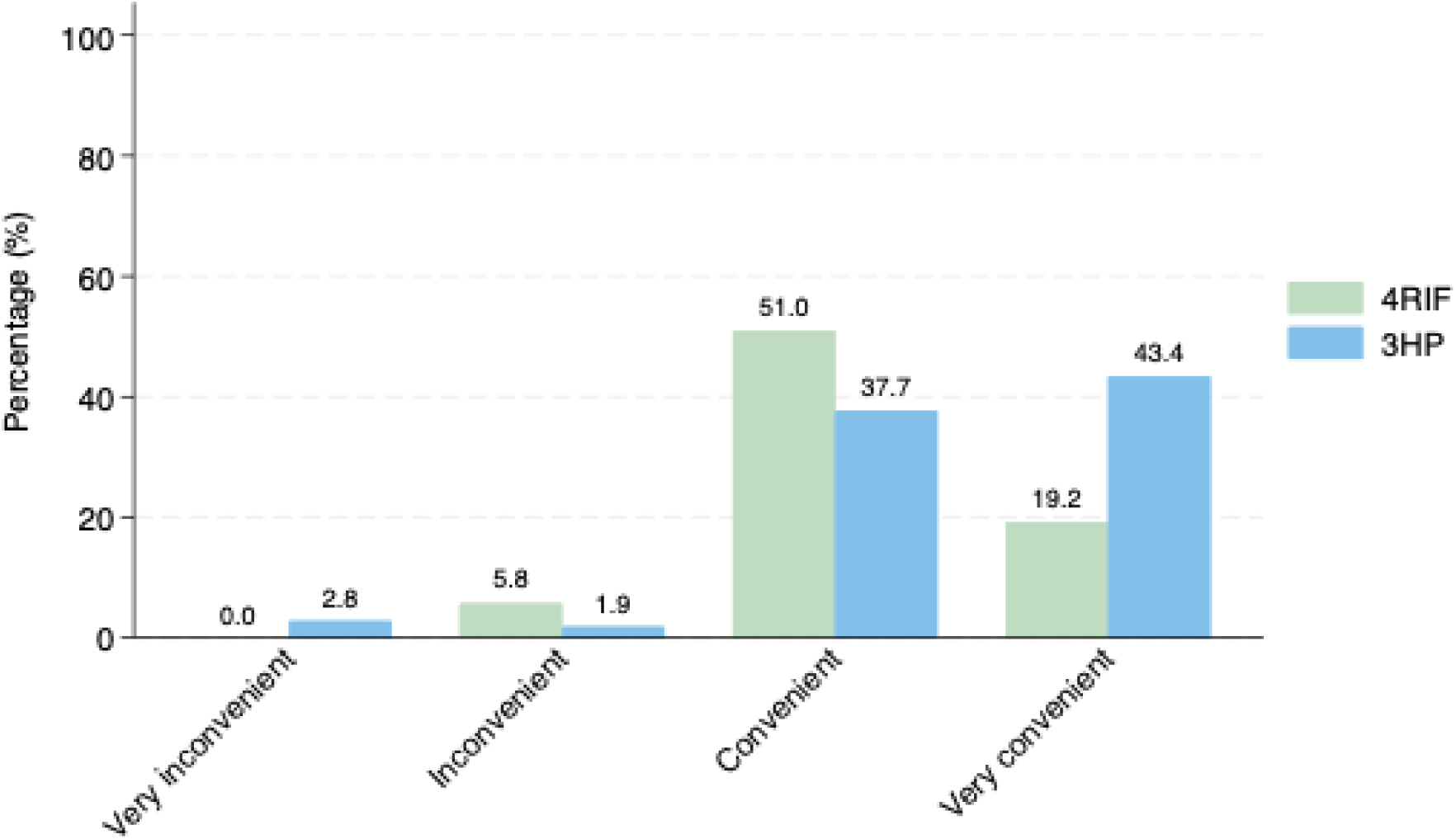
Bar graph of perceived convenience of the regimen by group

**Figure S4.**
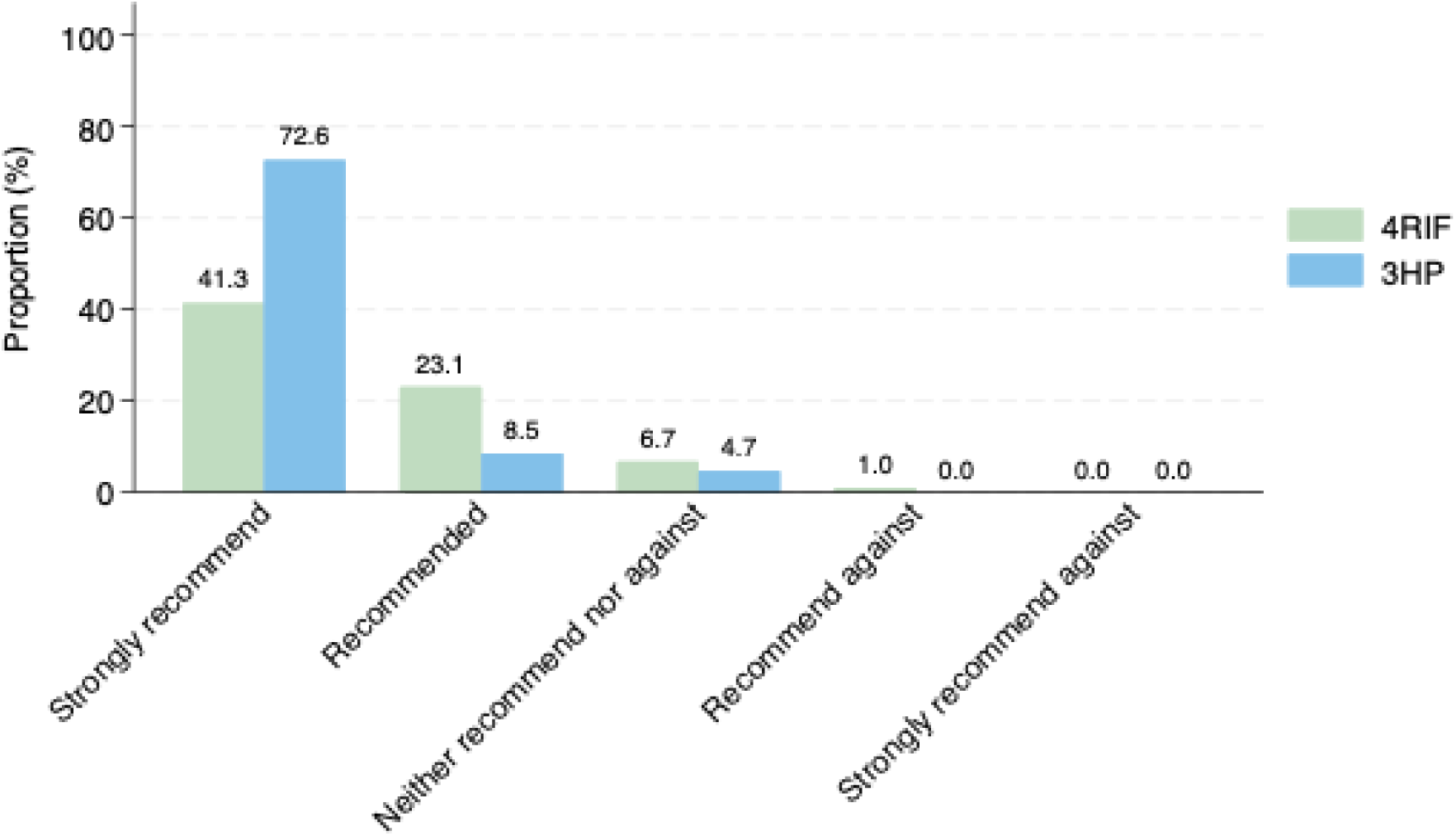
Bar graph of likelihood of recommending the regimen to others by group

**Figure S5.**
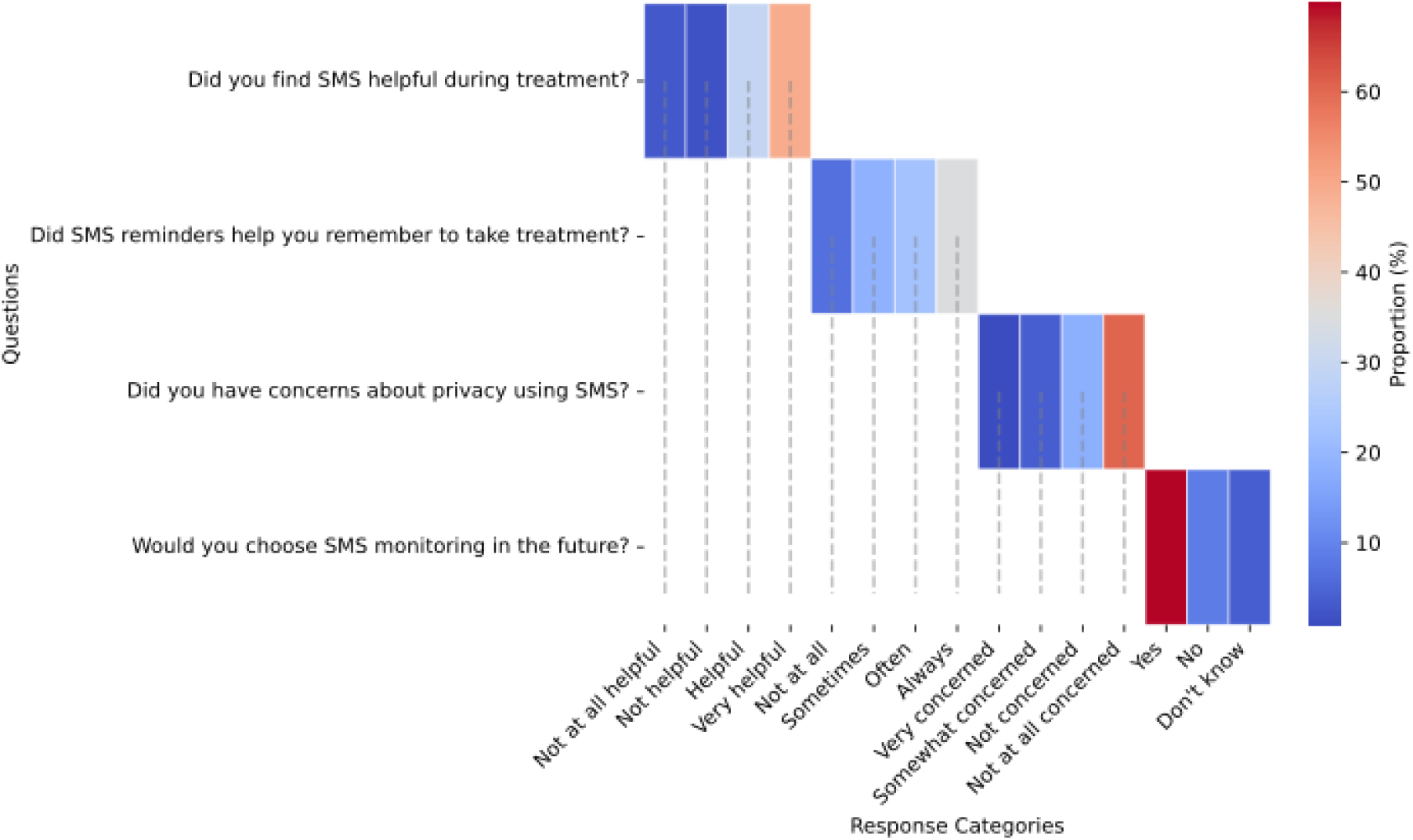
Heatmap of satisfaction with SMS reminders among participants taking 3HP regimen

## Supplementary Tables

**Table S1.** Weight-based weekly Rifapentine dosing.

| Weight | Weekly dose of rifapentine | Dose (mg/kg) |
| --- | --- | --- |
| 10-14kg | 300mg | 21.4-30.0 |
| 14.1-25kg | 450mg | 18.0-31.9 |
| 25.1kg-32kg | 600mg | 18.8-23.9 |
| 32.1kg-50kg | 750mg | 15.0-23.4 |
| >50kg | 900mg | ≤18.0 |

**Table S2.** Number of participants completed assigned treatments by group.

|  |  | **4RIF** | **3HP** | **Overall** |
| --- | --- | --- | --- | --- |
|  | **% of therapy taken** | **(n=104)** | **(n=106)** | **(n=210)** |
| **Therapy completed** | 100% | 55 (52.9) | 89 (84.0) | 144 (68.6) |
|  | ≥ 90 % | 13 (12.5) | 1 (0.9) | 14 (6.7) |
| **Therapy not completed** | Never commenced therapy | 20 (19.2) | 12 (11.3) | 32 (15.2) |
|  | Completed <10% | 4 (3.9) | 1 (0.9) | 5 (2.4) |
|  | 10-19% | 1 (1.0) | 1 (0.9) | 2 (1.0) |
|  | 20-29% | 4 (3.9) | 1 (0.9) | 5 (2.4) |
|  | 30-39% | 0 | 0 | 0 |
|  | 40-49% | 0 | 0 | 0 |
|  | 50-59% | 2 (1.9) | 0 | 2 (1.0) |
|  | 60-69% | 1 (1.0) | 1 (0.9) | 2 (1.0) |
|  | 70-79% | 3 (2.9) | 0 | 3 (1.4) |
|  | 80-89% | 1 (1.0) | 0 | 1 (0.5) |

**Table S3.** Patient satisfaction with assigned regimen by group (ITT population)

|  | **Response Categories** | **4RIF**<br>**(n=104)** | **3HP**<br>**(n=106)** | **Overall**<br>**(n=210)** |
| --- | --- | --- | --- | --- |
| **Convenience**<br>**rating** | Very inconvenient | 0 (0%) | 3 (2.8%) | 3 (1.4%) |
|  | Inconvenient | 6 (5.8%) | 2 (1.9%) | 5 (2.4%) |
|  | Convenient | 53 (51.0%) | 40 (37.7%) | 93 (44.3%) |
|  | Very convenient | 20 (19.2%) | 46 (43.4%) | 66 (31.4%) |
|  | Missing | 25 (24.0%) | 15 (14.2%) | 40 (19.0%) |
| **Recommend**<br>**LTBI**<br>**treatment to**<br>**others** | Strongly recommend | 43 (41.4%) | 77 (72.6%) | 120 (57.1%) |
|  | Recommended | 24 (23.1%) | 9 (8.5%) | 33 (15.7%) |
|  | Neither recommend for nor against | 7 (6.7%) | 5 (4.7%) | 12 (5.7%) |
|  | Recommend against | 1 (1.0%) | 0 (0%) | 1 (0.5%) |
|  | Strongly recommend against | 0 (0%) | 0 (0%) | 0 (0%) |
|  | Missing | 29 (27.9%) | 15 (14.2%) | 44 (21.0%) |

**Table S4.** Patient satisfaction with weekly SMS adherence reminders in 3HP treatment group (ITT population)

| Questions | Response Categories | Patients (n=106) |
| --- | --- | --- |
| Did you find SMS helpful during your treatment? | Not at all helpful | 3 (2.8%) |
|  | Not helpful | 2 (1.9%) |
|  | Helpful | 31 (29.3%) |
|  | Very helpful | 52 (49.1%) |
|  | Missing | 18 (17.0%) |
| Did SMS reminders help you remember to take treatment? | Not at all | 7 (6.6%) |
|  | Sometimes | 20 (18.9%) |
|  | Often | 24 (22.6%) |
|  | Always | 37 (34.9%) |
|  | Missing | 18 (17.0%) |
| Did you have any concerns about breaching your privacy by using SMS? | Very concerned | 1 (0.9%) |
|  | Somewhat concerned | 4 (3.8%) |
|  | Not concerned | 19 (17.9%) |
|  | Not at all concerned | 64 (60.4%) |
|  | Missing | 18 (17.0%) |
| Would you choose SMS monitoring in future? | Yes | 74 (69.8%) |
|  | No | 9 (8.5%) |
|  | Don't know | 4 (3.8%) |
|  | Missing | 19 (17.9%) |

**Table S5.** Types and grading of adverse events by group.

|  | 4RIF<br>(n=21*) |  |  |  | 3HP<br>(n=29*) |  |  |  |
| --- | --- | --- | --- | --- | --- | --- | --- | --- |
|  | Grading of AE (1-4) |  |  |  |  |  |  |  |
| Type pf AE | 1 | 2 | 3 | 4 | 1 | 2 | 3 | 4 |
| Rash | 1<br>(4.8) | 0 | 0 | 0 | 4<br>(13.8) | 0 | 0 | 0 |
| Headache | 3<br>(14.3) | 0 | 0 | 0 | 4<br>(13.8) | 0 | 0 | 0 |
| Gastrointestinal problem | 5<br>(23.8) | 1<br>(4.8) | 0 | 0 | 7<br>(24.1) | 1<br>(3.4) | 0 | 0 |
| Itchy skin | 0 | 0 | 0 | 0 | 2<br>(6.9) | 0 | 0 | 0 |
| Sleeping or mood problems | 2<br>(9.5) | 0 | 0 | 0 | 1<br>(3.4) | 0 | 0 | 0 |
| Liver toxicity / hepatitis | 2<br>(9.5) | 1<br>(4.8) | 0 | 0 | 1<br>(3.4) | 0 | 0 | 0 |
| Pregnancy | 2<br>(9.5) | 0 | 0 | 0 | 0 | 0 | 0 | 0 |
| Haematological abnormality | 0 | 0 | 0 | 0 | 1<br>(3.4) | 0 | 0 | 0 |
| Neuropathy | 0 | 0 | 0 | 0 | 2<br>(6.9) | 0 | 0 | 0 |
| Flu-like symptoms | 2<br>(9.5) | 0 | 0 | 0 | 6<br>(20.7) | 0 | 0 | 0 |
| Others (hair loss) | 1<br>(4.8) | 0 | 0 | 0 | 0 | 0 | 0 | 0 |
| Systemic drug reaction/hypersensitivity reaction * | 0 | 0 | 1<br>(4.8) | 0 | 0 | 0 | 0 | 0 |
| Total no. of AEs, n | 18<br>(85.7) | 2<br>(9.5) | 1<br>(4.8) | 0 | 28<br>(96.6) | 1<br>(3.4) | 0 | 0 |
|  | 4RIF<br>(n=104^) |  |  |  | 3HP<br>(n=106^) |  |  |  |
| Participants reported any AEs, n (%) | 18<br>(17.3) | 2<br>(1.9) | 1<br>(1.0) | 0 | 25<br>(23.6) | 1<br>(0.9) | 0 | 0 |

\*Total number of AEs in each treatment group. ^Total number of participants in each treatment group.

**Table S6.** Treatment decision for patients reported any AEs by group (ITT population)

|  | Treatment continued |  | Treatment stopped |  |  |  |
| --- | --- | --- | --- | --- | --- | --- |
|  |  |  | Patient's decision |  | Medical officer's decision |  |
|  | 4RIF<br>(n=104) | 3HP<br>(n=106) | 4RIF<br>(n=104) | 3HP<br>(n=106) | 4RIF<br>(n=104) | 3HP<br>(n=106) |
| Rash | 1 (1.0) | 4 (3.8) | 0 | 0 | 0 | 0 |
| Headache | 2 (1.9) | 4 (3.8) | 1 (1.0) | 0 | 0 | 0 |
| Gastrointestinal problem | 3 (2.9) | 7 (6.6) | 3 (2.9) | 1 (0.9) | 0 | 0 |
| Itchy skin | 0 | 2 (1.9) | 0 | 0 | 0 | 0 |
| Sleeping or mood problems | 2 (1.9) | 1 (0.9) | 0 | 0 | 0 | 0 |
| Liver toxicity / hepatitis | 0 | 1 (0.9) | 3 (2.9) | 0 | 0 | 0 |
| Pregnancy | 0 | 0 | 0 | 0 | 2 (1.9) | 0 |
| Haematological abnormality | 0 | 0 | 0 | 1 (0.9) | 0 | 0 |
| Neuropathy | 0 | 0 | 0 | 2 (1.9) | 0 | 0 |
| Flu-like symptoms | 1 (1.0) | 6 (5.7) | 1 (1.0) | 0 | 0 | 0 |
| Others (hair loss) | 0 | 0 | 1 (1.0) | 0 | 0 | 0 |
| Systemic drug reaction/hypersensitivity reaction * | 0 | 0 | 0 | 0 | 1 (1.0) | 0 |
| **Overall** | 9 (8.7) | 22 (20.8) | 9 (8.7) | 4 (3.8) | 3 (2.9) | 0 |

\* The definition of a systemic drug reaction followed the definition used in the PREVENT TB trial [11]. The PREVENT Tuberculosis sub-study protocol team established two broad criteria for such a drug reaction; (1) hypotension (systolic blood pressure <90 mm Hg), urticaria (hives), angioedema, acute bronchospasm, or conjunctivitis (red eyes); and (2) at least 4 of the following symptoms occurring concurrently (>1 of which had to be grade 2 or higher): weakness, fatigue, nausea, vomiting, headache, fever, aches, sweats, dizziness, shortness of breath, flushing, or chills. AE that met either of the above criteria were termed systemic drug reactions (SDR).

**Table S7.** Adjusted analysis for the primary outcome (ITT population)

|  | **4RIF**<br>**(N=104)** | **3HP**<br>**(N=106)** | **aRR\*** | **95% CI** | **p** |
| --- | --- | --- | --- | --- | --- |
| Proportion of participants completing treatment, n (%) | 68 (65.4%) | 90 (84.9%) | 1.29 | 1.10, 1.51 | 0.002 |

\*Adjusted relative risk, accounting for age and sex. Based on the assessment of baseline characteristics, no obvious imbalance was observed, suggesting minimal confounding; therefore, no additional covariates were adjusted. The adjusted relative risk closely aligns with the unadjusted results, demonstrating the robustness of the findings. In addition, there were no missing values for the primary outcome or covariates, thus imputation was not required.

**Table S8.** Adjusted analysis for the secondary outcomes (ITT population)

| Secondary outcomes | aOR* | 95% CI | p |
| --- | --- | --- | --- |
| Perceived convenience of the LTBI regimen | 2.68 | 1.43, 5.02 | 0.002 |
| Likelihood of recommending the assigned regimen to others | 3.84 | 1.82, 8.07 | <0.001 |

\*Adjusted for age and sex. The adjusted ORs, accounting for age and sex, closely aligned with the unadjusted results. Since there were no missing values for the covariates, imputation was not performed. See Table S3 for category-level survey responses (number and percentage).

**Table S9.**
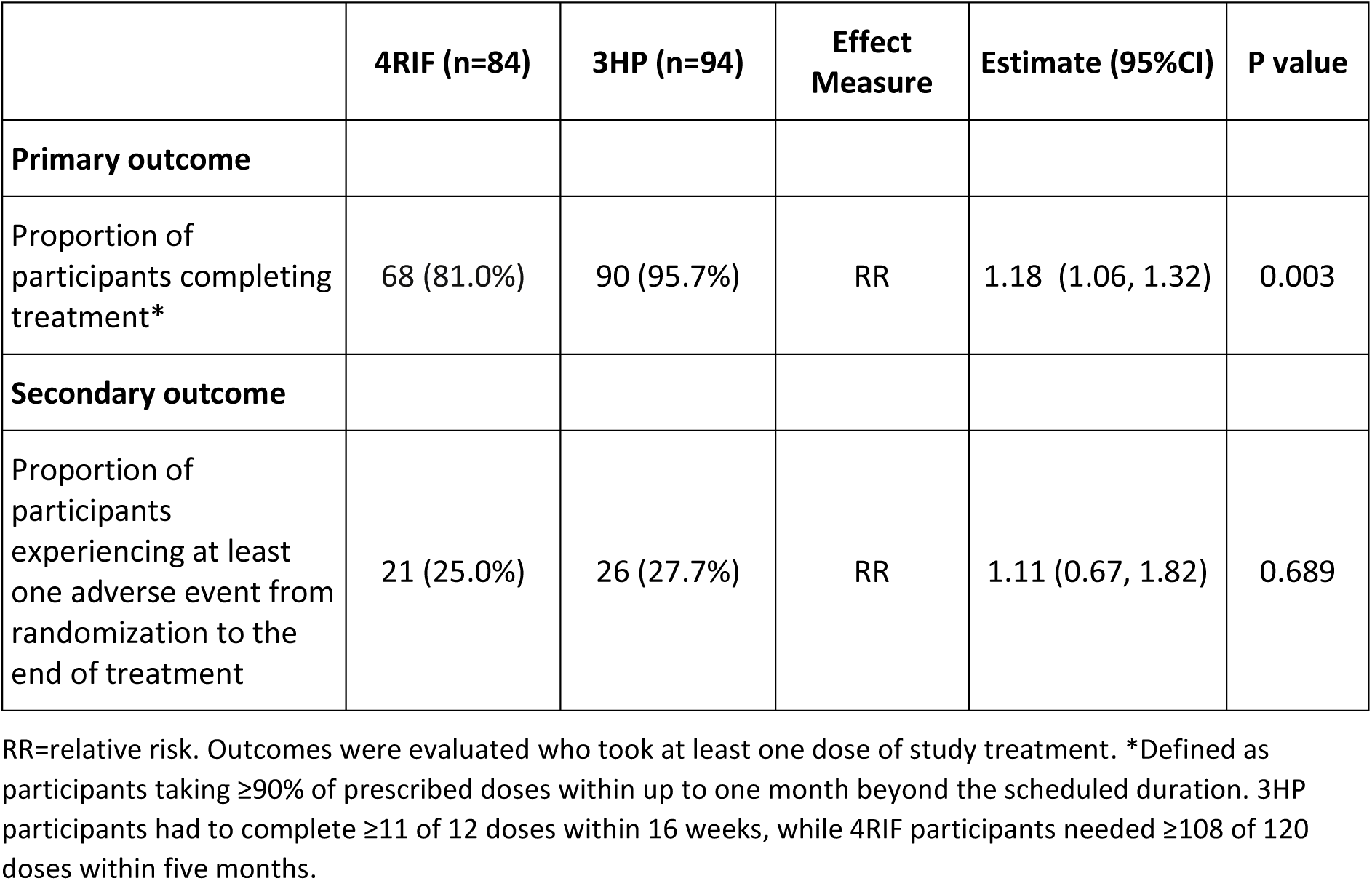
Sensitivity analyses showing outcomes among participants who took one or more doses of treatment.

**Table S10.** CONSORT checklist of information to include when reporting a randomised trial.

| Section/Topic | Item No | Checklist item | Reported on page No |
| --- | --- | --- | --- |
| **Title and abstract** |  |  |  |
|  | 1a | Identification as a randomised trial in the title | 1 |
|  | 1b | Structured summary of trial design, methods, results, and conclusions (for specific guidance see CONSORT for abstracts) | 1,2 |
| **Introduction** |  |  |  |
| Background and objectives | 2a | Scientific background and explanation of rationale | 3-5 |
|  | 2b | Specific objectives or hypotheses | 7 |
| **Methods** |  |  |  |
| Trial design | 3a | Description of trial design (such as parallel, factorial) including allocation ratio | 5, 6 |
|  | 3b | Important changes to methods after trial commencement (such as eligibility criteria), with reasons | NA |
| Participants | 4a | Eligibility criteria for participants | 5, 6 |
|  | 4b | Settings and locations where the data were collected | 5 |
| Interventions | 5 | The interventions for each group with sufficient details to allow replication, including how and when they were actually administered | 6 |
| Outcomes | 6a | Completely defined pre-specified primary and secondary outcome measures, including how and when they were assessed | 7, 8 |
|  | 6b | Any changes to trial outcomes after the trial commenced, with reasons | NA |
| Sample size | 7a | How sample size was determined | 9 |
|  | 7b | When applicable, explanation of any interim analyses and stopping guidelines | NA |
| Randomisation: |  |  |  |
| Sequence generation | 8a | Method used to generate the random allocation sequence | 6 |
|  | 8b | Type of randomisation; details of any restriction (such as blocking and block size) | 6 |
| Allocation concealment mechanism | 9 | Mechanism used to implement the random allocation sequence (such as sequentially numbered containers), describing any steps taken to conceal the sequence until interventions were assigned | 6 |
| Implementation | 10 | Who generated the random allocation sequence, who enrolled participants, and who assigned participants to interventions | 6 |
| Blinding | 11a | If done, who was blinded after assignment to interventions (for example, participants, care providers, those assessing outcomes) and how | 6 |
|  | 11b | If relevant, description of the similarity of interventions | 6 |
| Statistical methods | 12a | Statistical methods used to compare groups for primary and secondary outcomes | 9, 10 |
|  | 12b | Methods for additional analyses, such as subgroup analyses and adjusted analyses | 9, 10 |
| **Results** |  |  |  |
| Participant flow (a diagram is strongly recommended) | 13a | For each group, the numbers of participants who were randomly assigned, received intended treatment, and were analysed for the primary outcome | 11 |
|  | 13b | For each group, losses and exclusions after randomisation, together with reasons | 11 |
| Recruitment | 14a | Dates defining the periods of recruitment and follow-up | 11 |
|  | 14b | Why the trial ended or was stopped | NA |
| Baseline data | 15 | A table showing baseline demographic and clinical characteristics for each group | 18 |
| Numbers analysed | 16 | For each group, number of participants (denominator) included in each analysis and whether the analysis was by original assigned groups | 11 |
| Outcomes and estimation | 17a | For each primary and secondary outcome, results for each group, and the estimated effect size and its precision (such as 95% confidence interval) | 11 |
|  | 17b | For binary outcomes, presentation of both absolute and relative effect sizes is recommended | 11, 21 |
| Ancillary analyses | 18 | Results of any other analyses performed, including subgroup analyses and adjusted analyses, distinguishing pre-specified from exploratory | 13, Supplementary 4-5 |
| Harms | 19 | All important harms or unintended effects in each group (for specific guidance see CONSORT for harms) | 12, 13, 22 |
| **Discussion** |  |  |  |
| Limitations | 20 | Trial limitations, addressing sources of potential bias, imprecision, and, if relevant, multiplicity of analyses | 13-17 |
| Generalisability | 21 | Generalisability (external validity, applicability) of the trial findings | 13-17 |
| Interpretation | 22 | Interpretation consistent with results, balancing benefits and harms, and considering other relevant evidence | 13-17 |
| **Other information** |  |  |  |
| Registration | 23 | Registration number and name of trial registry | 10 |
| Protocol | 24 | Where the full trial protocol can be accessed, if available | Supplementary 3 |
| Funding | 25 | Sources of funding and other support (such as supply of drugs), role of funders | 10 |

